# Comprehensive Management of High-Risk Populations for Stroke Based on Social Networks in China: A Multicenter Randomized Clinical Trial (COMPLIANCE-MT)

**DOI:** 10.64898/2026.01.23.26344106

**Authors:** Dongmei Li, Yu Zhou, Huanhuan Hu, Jing Zheng, Mengxia Chen, Mengting Qiao, Yanqiu Weng, Xiaoying Lu, Pengfei Yang, Jianmin Liu, Lingjuan Zhang, the Compliance-MT collaborators

**Affiliations:** Department of Cerebrovascular Center, Changhai Hospital, Naval Medical University, Shanghai, PR China; Nursing Department, Changhai Hospital, Naval Medical University, Shanghai, PR China; Nursing Teaching and Research Department, Changhai Hospital, Naval Medical University, Shanghai, PR China; Department of Nursing, the First Medical Center of Chinese PLA General Hospital, Beijing, PR China; Department of Neurology, the First Affiliated Hospital of Zhengzhou University, Zhengzhou, PR China

**Author notes:** **Corresponding author:** Dr. Lingjuan Zhang at Nursing Teaching and Research Department, Changhai Hospital, Naval Medical University, Shanghai, PR China. E; Dr. Pengfei Yang at Neurovascular Center, Changhai Hospital, Naval Medical University, Shanghai, China. E; Dr. Jianmin Liu at the Neurovascular Center, Changhai Hospital, Naval Medical University, Shanghai, China. E; Or Dr. Yu Zhou at the Neurovascular Center, Changhai Hospital, Naval Medical University, Shanghai, China. E. These authors contributed equally (Dongmei Li, Yu Zhou).

**Keywords:** Stroke, Medication Adherence, Digital Health Intervention, High-Risk P opulations, Primary Prevention, Secondary Prevention

## Abstract

**Background:** Suboptimal adherence to preventive medications and inadequate management of risk factors are major barriers to effective post-discharge care for individuals at high risk of stroke, particularly in resource-constrained settings. Digital health interventions delivered via widely accessible platforms offer a promising scalable approach. We conducted this trial to evaluate the efficacy of Social Network based patient care in improving medication adherence in high-risk populations for stroke.

**Method:** COMPLIANCE-MT is a multicenter, prospective, randomized, open-label, parallel-group trial with blinded outcome assessment (PROBE design) assessing the superiority of Social Network-based care versus conventional care in medication adherence for high-risk populations for stroke. A total of 720 participants will be recruited across 33 hospitals in China and randomized in a 1:1 ratio to either a 12-month Social Network (WeChat) based coordinated care program or routine follow-up. The primary outcome is the proportion of patients achieving more than 80% adherence to all indicated vascular prevention medications (antihypertensives, hypoglycemics, lipid-lowering agents, anticoagulants, and antiplatelets) at 12 months. Analysis will be performed on an intention-to-treat approach.

**Discussion:** The COMPLIANCE trial evaluates a social network-based intervention in improving adherence to five evidence-based vascular prevention medications in both primary prevention and secondary prevention. If proven effective, this model could inform national strategies for preventing strokes in resource-limited settings.

**Trial registration:** ClinicalTrials.gov NCT05963828.

## Introduction and rationale

Stroke remains a persistent and major global health challenge, accounting for millions of deaths and over 100 million disability-adjusted life years (DALYs) lost annually worldwide.^1^ Although evidence-based pharmacotherapy (antihypertensives, antidiabetics, lipid-lowering agents, anticoagulants, and antiplatelet drugs), risk factor control, and lifestyle modification—demonstrates significant efficacy in reducing stroke recurrence and associated morbidity.^2,3^ Approximately 40% of stroke survivors become non-adherent within three months after hospital discharge, which may place them at increased risk of stroke recurrence.^4^

Existing strategies, such as simple SMS reminders or resource-intensive community health worker programs, often face limitations in functional scope or scalability within resource-constrained settings. There is an urgent need for automated, low-cost delivery models that can facilitate complex, longitudinal care coordination without requiring significant infrastructure.^5^ The ubiquitous penetration of mobile social network platforms provides an unprecedented infrastructure for scalable health interventions. Unlike isolated mobile applications, social network-integrated programs leverage existing user habits, facilitating seamless multidisciplinary coordination between patients, caregivers, and specialists.^6–8^ Despite the potential for these digital ecosystems to enhance chronic disease management,^9,10^ robust clinical evidence for social network-mediated stroke care remains limited.

To address this critical evidence gap, we designed the Comprehensive Management of High-risk Populations for Stroke based on Social Networks in China: A Multicenter Randomized Clinical Trial (COMPLIANCE-MT) to evaluate the efficacy of a social network(WeChat)-integrated intervention improves medication adherence and risk factor control.

## Methods

### Design

COMPLIANCE-MT is a multicenter, prospective, parallel-group, randomized controlled superiority trial designed to evaluate the efficacy of a social network-based integrated management program compared with standard care for improving medication adherence and optimizing modifiable risk factor control in high-risk stroke populations during a 12-month follow-up period after discharge.(figure 1) Followed up would be conducted at 1, 3, 6, and 12 months after discharge.

**Figure 1.**
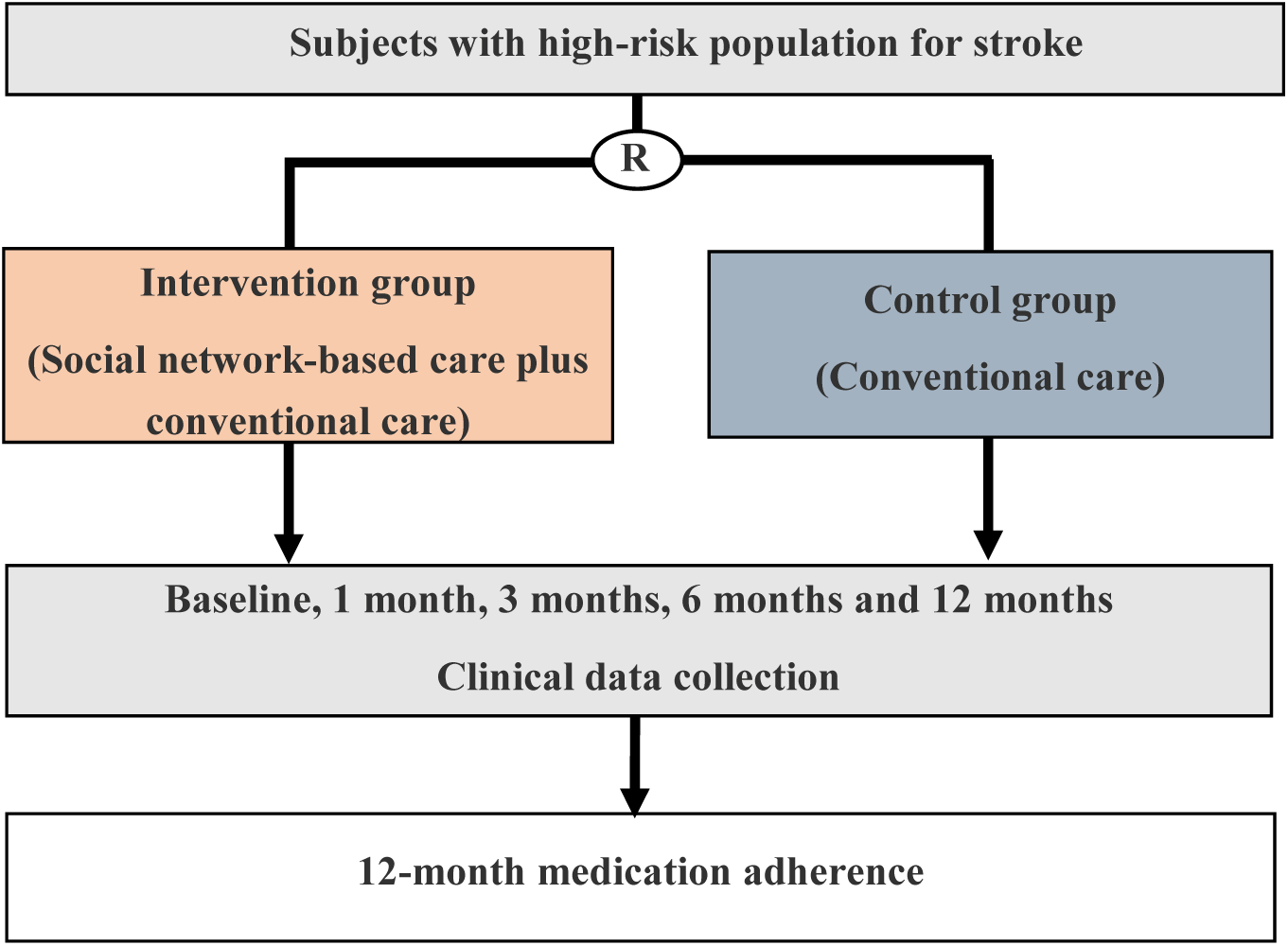
Randomized controlled trial design

Following the confirmation of participant eligibility according to pre-specified inclusion and exclusion criteria, formal informed consent will be obtained. All consenting procedures must be completed before the randomization process is initiated. The study is conducted in compliance with local and international regulatory and ethical requirements and was approved by was approved by ethics committee at each participating hospital prior any trial activities being undertaken.

### Patient inclusion and exclusion criteria

#### Inclusion criteria

1. Age ≥18 years.
2. Hospitalized patients at high risk of stroke are those who have at least three of the following risk factors: hypertension, dyslipidemia, diabetes, atrial fibrillation/valvular disease, smoking history, overweight/obesity, physical inactivity, or family stroke history; or those with a history of prior transient ischemic attack (TIA) or stroke.12
3. modified Rankin Scale (mRS) score ≤2.
4. Smartphone/WeChat access (patient or caregiver).
5. Informed consent obtained.
6. Active long-term therapy: ≥ 1 medication (antihypertensive, hypoglycemic, lipid-lowering, anticoagulant, antiplatelet).

#### Exclusion criteria

1. Inability to operate smartphones (patient or caregiver).
2. Comorbidities potentially confounding outcome assessments, including Advanced malignancies (life expectancy <12 months); Documented dementia; Severe psychiatric disorders (e.g., schizophrenia, major depressive disorder).
3. Residence in areas with unreliable internet access.
4. Concurrent participation in other clinical trials.
5. Any condition deemed by investigators to preclude safe trial participation.

### Randomization and blinding

Subjects will be randomized via an Internet-based randomization system in a 1:1 manner to treatment with social network-based intervention group or standard care group. Stratified randomization with permuted blocks will be performed to ensure balance across key prognostic factors, including clinical center and prevention Type (Primary vs. Secondary Prevention)

Both patients and treating physicians will be aware of the treatment assignment. But primary outcome evaluation would be blinded. Information about medication adherence will be accessed through standardized forms and procedures, by trained investigators blinded for treatment allocation.

An independent DSMB statistician will combine data on treatment allocation with the clinical data to report to the Data and Safety Monitoring Board (DSMB). The Steering Committee remains blinded to all analyses throughout the trial duration.

### Intervention

Eligible participants will be randomly assigned to either the conventional care group or social network-based intervention group.

Conventional care group: Patients in conventional care group will receive standardized education based on ASA/AHA 2021 guidelines prior to discharge,^2^ delivered verbally by a certified Brain-Heart Health Manager (BHHM) and supplemented with an expert-reviewed booklet. Content will cover medication adherence, risk factor control, stroke recognition, emergency response, and follow-up plans. A contact number will be provided for post-discharge support. A baseline archive will document demographics, lifestyle, and cardiovascular risk factors.

Social network-based intervention group: Prior to discharge, participants in Social network-based intervention group are onboard to the integrated digital platform in addition to conventional care. BHHMs facilitate the activation of the digital interface via a unique QR code, assist in the creation of a comprehensive electronic health record (EHR), and guide participants through an interactive tutorial to ensure technical proficiency in data entry and communication features. Post-discharge, participants in intervention group will receive comprehensive management via the social network-integrated digital platform, including pharmacotherapy support(automated, dose-specific push notifications for medication reminders and timestamped adherence verification), precision education(weekly multimedia content tailored to the participant’s dynamic risk profile, derived from AHA/ASA guidelines), biometric surveillance(real-time clinical alerts for abnormal biometric data[e.g., SBP >140mmHg]), telehealth access (on-demand secure messaging and video consultations with BHHMs, with a guaranteed response time within 24 hours.)

BHHMs conduct structured follow-ups at 1, 3, 6, and 12 months via telephone or clinic visits for participants in both groups. These sessions involve standardized assessments of medication adherence, physiological parameters, and psychological status, alongside individualized health counseling.

### Outcomes

#### Primary Outcome

The primary outcome is good Medication Adherence to all guideline-recommended vascular prevention medications at 12 months post-discharge

Adherence is assessed using self-reported data, with participants asked to indicate the number of days they missed taking a dose for each medication class during the preceding 30 days. This evaluation is conducted separately for each of the five evidence-based secondary prevention drug classes: antihypertensives, hypoglycemics, lipid-lowering agents, anticoagulants, and antiplatelets. Good adherence for each class is defined as taking the prescribed medication on more than 24 days out of the previous 30 days (corresponding to an adherence rate >80%). To meet the primary endpoint, a participant must achieve this >80% adherence threshold simultaneously across all five medication classes at the 12-month follow-up.

Any self-directed cessation or adjustment of the regimen without medical consultation is categorized as non-adherence. Conversely, patients who cease or adjust their medications according to medical advice will be considered adherent. For those who adjust their medications based on medical advice, adherence will be further assessed by inquiring about the number of days the medications were taken in the past month; if the number of days exceeds 24, they will be classified as adherent.

#### Secondary outcomes

1. Proportion of good medication adherence to stroke prevention drugs at 1-, 3-, 6-, and 12-months post-discharge (assessed using the Morisky-8 Medication Adherence Scale [MMAS-8]). Good adherence is defined as an MMAS-8 score greater than 6.^11^
2. Risk factor control, including blood glucose, blood pressure, lipid profile, body mass index (BMI), waist circumference, hip circumference, and smoking status, at 1-, 3-, 6-, and 12-months post-discharge.
3. Health-related quality of life (HRQoL) assessed using the EuroQol Five-Dimension Five-Level Scale (EQ-5D-5L) at 1-, 3-, 6-, and 12-months post-discharge.^12^
4. Anxiety symptom severity assessed using the 7-item Generalized Anxiety Disorder Scale (GAD-7) at 1-, 3-, 6-, and 12-months post-discharge.^13^
5. Depressive symptom severity assessed using the 9-item Patient Health Questionnaire (PHQ-9) at 1-, 3-, 6-, and 12-months post-discharge.^14^
6. Stroke prevention knowledge scores assessed using the Stroke Prevention Knowledge Questionnaire at 1-, 3-, 6-, and 12-months post-discharge.^15^
7. Personal motivation for stroke prevention assessed using the Stroke Attitude Questionnaire at 1-, 3-, 6-, and 12-months post-discharge.^16^
8. Perceived social support assessed using the Perceived Social Support Scale (PSSS) at 1-, 3-, 6-, and 12-months post-discharge.^17^
9. Stroke prevention-related health behavior scores assessed using the Stroke Prevention Health Behavior Scale at 1-, 3-, 6-, and 12-months post-discharge.^18^
10. Self-efficacy for chronic disease management assessed using the Chronic Disease Self-Efficacy Scale at 1-, 3-, 6-, and 12-months post-discharge.^19^
11. Intentions regarding prehospital delay in stroke emergency care assessed using the Prehospital Delay Behavior Intention Scale for Stroke at 1-, 3-, 6-, and 12-months post-discharge.^20^
12. Incidence of major adverse cerebrovascular and cardiovascular events (MACCE), including stroke, acute coronary syndrome, and vascular death, at 1-, 3-, 6-, and 12-months post-discharge.

### Data collection and management

Data will be entered by clinical investigators and supporting trial personnel. Primary outcome data were obtained from interviews that were performed in person or by telephone conducted by trained physicians who were unaware of the trial-group assignments. During interview, participants were asked to indicate the number of days they missed taking a dose for each medication class during the preceding 30 days. Other clinical assessments at baseline, at 1-, 3-, 6-, and 12-months post-discharge were performed by BHHMs. All the clinician assessors received both on-site training on how to deliver patient education and perform the clinical assessments.

### Data and Safety Monitoring Board

An independent Data and Safety Monitoring Board (DSMB) oversees the study’s safety, ethics, and accumulating data. Governed by a charter, the DSMB monitors efficacy and safety outcomes for early identification of significant benefits or potential harms. They provide recommendations to the Trial Steering Committee (TSC) on whether to continue, pause, or terminate recruitment based on unblinded data reviews of recruitment, protocol adherence, primary and secondary outcomes, and serious adverse events (SAEs) at regular intervals.

### Sample size estimates

The sample size was determined based on a pre-trial feasibility survey involving 3702 similar patients across 26 centers in China, which revealed a composite medication adherence rate of 77.3% among high-risk stroke patients under conventional care.We hypothesized a clinically meaningful absolute improvement of 10% in adherence rates with a social network-based coordinated care intervention (from 77.3% in the control group to 87.3% in the intervention group), an effect size consistent with recent cardiovascular digital health trials showing moderate improvements in adherence through remote or community-based strategies.^21,22^ Using PASS 11.0 software and Fisher’s exact test, a sample of 648 patients (324 per group) was calculated to provide 90% power to detect this difference at a two-sided significance level of 0.05. To account for an anticipated 10% attrition rate over the 12-month follow-up in this high-risk stroke population, the target recruitment was inflated to 720 participants (360 per group).

To ensure the study’s robustness, a sensitivity analysis was performed. This analysis confirmed that even if the control group adherence rate is lower than our survey’s findings (ranging from 65.1% to 71.6%, as reported in previous literature), a sample size of 720 remains sufficient to detect a 10% improvement with a statistical power between 84% and 89%.

### Statistical analysis

All analyses will be conducted on an intention-to-treat basis. Baseline characteristics will be summarized by management group, and missing baseline data will be reported. Missing baseline values will be imputed using regression imputation.

The primary endpoint will be analyzed by means of a binary logistic regression. Subgroups will be analyzed by testing for interaction between specific baseline characteristics and treatment Pre-defined Statistical analyses will be detailed in the pre-specified Statistical Analysis Plan. Secondary outcome analyses will be considered exploratory. The main analysis for these endpoints will utilize a complete case approach under the assumption of a missing-completely-at-random (MCAR) mechanism.

### Progress to date

Patient recruitment was initiated on July 19, 2023, across participating centers in China and successfully concluded on March 29, 2024. The 12-month longitudinal follow-up phase was completed on April 30, 2025. Currently, the trial database has been locked, and the Statistical Analysis Plan (SAP) is being finalized prior to unblinding and formal data analysis.

## Discussion

The present study introduces a social network-based digital management platform that represents a significant advance over existing models for stroke prevention, with distinct innovations compared to traditional text message (SMS) interventions and community health worker-led programs. Conventional SMS-based interventions are predominantly unidirectional, focusing solely on medication reminders without feedback loops or personalized adjustments, leading to inconsistent long-term adherence and clinical outcomes.^23,24^ In contrast, the WeChat-based platform establishes a closed-loop, multi-dimensional ecosystem that integrates the full continuum of care—from initial risk stratification and real-time health monitoring to tailored education, on-demand multidisciplinary consultation, and structured follow-up. This design addresses the passivity of one-way messaging by enabling dynamic bidirectional communication: patients can actively report health data, seek clarification on medication use, and receive personalized feedback, while clinicians can promptly respond to abnormal indicators and adjust management plans. Compared to community-based programs that rely on community health workers, our intervention overcomes scalability barriers—community health worker models require substantial human resources for face-to-face interactions, limiting their reach in resource-constrained settings,^25^ whereas the digital platform allows simultaneous management of large cohorts with standardized protocols and minimal marginal costs.^26^

The primary endpoint—medication adherence—was defined to optimize clinical relevance, statistical power, and methodological objectivity. While a universal gold standard for adherence measurement remains elusive, we adopted a threshold of >=80% (defined as taking prescribed medications for at least 24 out of 30 days) for five core evidence-based vascular prevention classes: antihypertensives, hypoglycemics, lipid-lowering agents, anticoagulants, and antiplatelets. Although the study initially considered the Morisky Medication Adherence Scale (MMAS) as the primary outcome measure, we ultimately changed the primary endpoint to the Composite Medication Adherence Score (CMAS). While the MMAS provides a subjective, questionnaire-based assessment of adherence behavior, the CMAS yields a discrete, objective numerical value representing medication persistence (typically calculated as the proportion of days covered across multiple medication classes). This makes the CMAS more directly comparable across different pharmacotherapeutic agents. By adopting a binary threshold of >80% based on actual days of medication availability, we enhance the objectivity of the primary endpoint and better align our study with the rigorous standards commonly used in contemporary cardiovascular outcome trials. Beyond adherence, the comprehensive suite of secondary outcomes captures physiological control (blood pressure, glucose, lipids), patient-reported outcomes (quality of life via EQ-5D-5L; psychological status via GAD-7/PHQ-9), health behaviors, and major adverse cerebrovascular/cardiovascular events, addressing the narrow focus on single endpoints seen in some prior research. ^27^

By utilizing a low-barrier interface (no separate application download, large-font options, and multimedia-based education), the platform minimizes the “digital divide” common among older stroke survivors. This user-centric design ensures that technological complexity does not become a barrier to care, particularly for patients with post-stroke functional or cognitive impairments.

Building upon prior evidence for digital health interventions in cardiovascular disease,^3,26^ this trial innovates by integrating social connectivity, structured multidisciplinary support, and standardized care delivery into a scalable model via a pre-existing, widely adopted social platform. If proven effective, this approach could inform sustainable, resource-efficient strategies for integrated stroke prevention.

## Data Availability

All data produced in the present study are available upon reasonable request to the authors

